# Prevalence and associated factors of pterygium among adults living in Kolla diba town, Northwest Ethiopia

**DOI:** 10.1101/2019.12.11.19014290

**Authors:** Tibebu Kassie Alemayehu, Yezinash Addis, Zewdu Yenegeta Bizuneh, Mebratu Mulusew Tegegne, Abiy Maru Alemayehu

**Affiliations:** Department of Optometry, School of Medicine, College of Medicine Health Science, University of Gondar, Gondar city, Ethiopia

**Author notes:** Tibebu Kassie Alemayehu, (TK). These authors contributed equally to this work (Yezinash Addis, Zewdu Yenegeta Bizuneh, Mebratu Mulusew Tegegne, Abiy Maru Alemayehu).

**Keywords:** Pterygium, Marital status, Prevalence, Kolla Diba, Ethiopia

## Abstract

**Introduction:** Pterygium is a disfiguring disease that can potentially lead to blindness and has a significant public health problem in rural areas. It is more common in warm, windy and dry climates of tropical and sub-tropical “pterygium belt” regions of Africa, including Ethiopia. Globally, the prevalence ranging from 0.07% to 53%. Studies conducted on the prevalence of pterygium in developing countries like Ethiopia were limited with wider discrepancy between them.

**Objective:** The aim of this study was to assess the prevalence of pterygium and associated factors among adults in Kolla Diba town, Northwest Ethiopia, 2019.

**Method:** A community-based cross-sectional study was done in Kolla Diba town from May 30-June 16, 2019. Systematic random sampling technique was used to select 627 study participants. Data was collected through basic ophthalmic examination with portable slit lamp, 3x magnifying loop and torchlight and a pretested structured questionnaire was administered to collect the data. Then the data entered into EPI INFO version 7 and analyzed using SPSS version 20. Descriptive statistics and binary logistic regression analysis were employed. P-values of <0.05 was considered statistically significant.

**Result:** A total of 605 study participants were involved with a response rate of 96.5%. Among them 317 (52.4%) participants were males. The mean age of the respondents was 38.18 ± 15.56 with a range of (18-95) in years. The overall Prevalence of pterygium among adults living in Kolla Diba town was 112 (18.5% (95% CI (15.6-21.7)). Being widowed (AOR = 7.32 (95%CI: 2.88, 18.57)), outdoor occupation (AOR = 2.50 (95%CI: 1.46, 4.29)), sun exposure (AOR = 2.38 (95%CI: 1.28, 4.43)), wind exposure (AOR = 1.97 (95%CI: 1.04, 3.72)), alcohol drinking (AOR = 2.26 (95%CI: 1.48, 4.63)), and severe blepharitis (AOR = 2.45 (95%CI: 1.48, 4.05)) had statistically significant positive association with pterygium.

**Conclusion:** The prevalence of pterygium was relatively higher. Marital status (being widowed), outdoor occupation, sun exposure, wind exposure, alcohol drinking, and severe blepharitis were significantly associated with the development of pterygium.

## Introduction

Pterygium is a benign, radially oriented, fleshy triangular/wingy shaped abnormal fibro-vascular growth of the conjunctiva encroaching towards the central cornea in either one or both eyes [1,2]. The main clinical presentations are redness, irritation, foreign body sensation, decreased vision and ocular discomfort [3,4]. The severity of pterygium can be graded as grade 1, 2, 3 and 4 based on the location of pterygium head encroachment on the cornea [5,6]. Genetics and other environmental factors such as wind, dry atmosphere, dust, chemicals, air pollution, geographical location, and hereditary factors may contribute for the development of pterygium [7]. Pterygium is more common in warm, windy and dry climates of tropical and subtropical regions [8,9]. The burden of pterygium is higher among individuals who live in rural residence, low economic status, illiterates, and outdoor workers, who spend a greater part of the day outdoor under excessive sun exposure, wind, and dust [10–13].

Globally, the prevalence of pterygium ranges from 0.07% to 53% worldwide and 8.0% to 38.7% in Africa [2,3,6,12,14,15]. In Ethiopia, the prevalence was 8.8% in Meskan district, Southern Ethiopia and 38.7% in Gondar city, Northwest Ethiopia [2,16].

Untreated pterygium may lead to visual impairment, cosmetic problems, restriction of ocular motility, significant socioeconomic problems and reduced quality of life [17–21]. Even though, the consequence is higher, it is possible to minimize the chances of being afflicted with pterygium by wearing UV protective sun glasses, wide-brimmed hats, caps and using umbrella to reduce the exposure to environmental irritants [2,6,10,15,22,23].

Pterygium is a common ophthalmic condition in “pterygium belt” regions, located between latitudes 40° N and 40 ° S of the equator [7,15,24]. The exposure to environmental factors is also higher in Africa including Ethiopia but there is a scarcity of evidence on the prevalence of pterygium in this region [13,25–29]. Therefore, this study aimed to determine the prevalence and associated factors of pterygium among adults in Kolla Diba town. The result of this study will be important for planning and implementing health care services and also act as base line data for further studies.

## Methods and Materials

### Study design, setting and sampling

A community-based cross-sectional study was conducted in Kolla Diba town, Northwest Ethiopia, located 712km from the capital city of Ethiopia, Addis Ababa. The altitude of the town is 1827m above sea level. According to statistical data from Dembiya woreda Administrative office, Kolla Diba town has a total population of 25,831 with three kebeles including 5328 households. The total number of adults (≥ 18 years) living in Kolla Diba town was 15,534. In the town, there is one primary hospital and one health center.

A total of 627 sample size was determined by single proportional formula by considering 10% non-response rate. In the study, 605 study participants were recruited and completed the questionnaire along with basic ophthalmic examinations with a response rate of 96.5%. Households from all kebeles were chosen by using systematic random sampling technique with a sampling fraction of eight. The first household was selected by lottery method between, the 1^st^ & 8^th^ household and then continued in every 8^th^ household. Finally, one adult in each participating household with age ≥ 18 years old was randomly selected and recruited as study participant.

The study was conducted in accordance with the Declaration of Helsinki and approved by the University of Gondar Ethical Review Board. In accordance with the Ethiopian National Research Ethics Review Guideline, verbal informed consent was obtained from all adults ≥18 years old using an information sheet in the local language “Amharic”. Since the study didn’t involve invasive eye examination procedures, the university ethical review board approved verbal informed consent. The study participants’ agreements were first obtained verbally prior to data collection. Then the data was collected by trained senior optometrists. Those adults who had pterygium were prescribed sunglass and referred to the University of Gondar tertiary eye care and training center for further examination and management.

### Operational definition

Sun exposure divided into two groups by crude cut off value of sun exposure duration: a participant who exposed to sunlight for above or equal to 5hrs per day considered as exposed, whereas less than 5hrs per day considered as non-exposed [26].

### Grading of pterygium

Grade 1: head of a pterygium at the limbus

Grade 2: head of a pterygium between the limbus and the un-dilated pupil margin Grade 3: head of a pterygium at the pupil margin

Grade 4: head of a pterygium within the pupil margin [35]

## Data collection

The pre-tested and structured questionnaire of local language ‘Amharic’ was used to carry out interview with adults’ ≥18 years old <u>(S1 Questionnaire).</u> The study tools were pretested in a nearby area of study site that had not been sampled (Chuahit), on 5% of the sample who fulfilled the sampling inclusion criteria, to validate questions and observations. The collected data from study participants include information’s on socio-demographic characteristic, behavioral/ lifestyle related factors and environmental factors. Face to face interview by using standardized questionnaires were conducted on those selected participants. Standard basic ophthalmic examination by using portable slit lamp, torch light and 3x magnifying loop were done by trained senior optometrists in order to rule out the diagnosis of pterygium. On the fieldwork, the quality of data was cross checked (5% of the filled questionnaire) for completeness, accuracy, and clarity through supervision on daily basis.

### Statistical analysis

After coding, the data was entered into EPI INFO 7, exported and analyzed by using SPSS version 20. The descriptive statistics were carried out to compute different proportions and summary statistics. Model fitness was checked by using the Hosmer and Lemeshow. Variables were fitted into the model by using the backward stepwise method. The analytical statistics was done by using bivariate and multivariate logistic regression. Those variables with 95% CI and p-value less than 0.05 were considered as statistically significant factors of pterygium.

## Results

A total of 605 participants were recruited and completed the questionnaire along with basic ophthalmic examinations with a response rate of 96.5%. Among them 317 (52.4%) participants were males. The mean age of the respondents was 38.18 ± 15.56 with a range of (18-95) in years. Majority 188 (31.1%) of the respondents completed secondary school and almost half of the participants got married 303 (50.1%). Most of the participants were indoor workers 410 (67.8%). Majority of the participants had an income of ≤ 1000 ETB from monthly median income of 2075.50 ETB. <u>Table 1</u>.

**Table 1:** Socio-demographic characteristics among adults living in Kolla Diba town, Northwest Ethiopia, June 2019 (n=605).

| **Characteristics** | **Frequency** | **Percent (%)** |
| --- | --- | --- |
| **Age (in years)** |  |  |
| 18-26 | 169 | 27.9 |
| 27-35 | 161 | 26.6 |
| 36-48 | 133 | 22.0 |
| ≥49 | 142 | 23.5 |
| **Gender** |  |  |
| Male | 317 | 52.4 |
| Female | 288 | 47.6 |
| **Marital status** |  |  |
| Single | 179 | 29.6 |
| Married | 303 | 50.1 |
| Divorced | 80 | 13.2 |
| Widowed | 43 | 7.1 |
| **Educational status** |  |  |
| Unable to read and write | 99 | 16.4 |
| Read & write only | 80 | 13.2 |
| Primary school | 100 | 16.5 |
| Secondary school | 188 | 31.1 |
| College and above | 138 | 22.8 |
| **Type of occupation** |  |  |
| Indoor* | 410 | 67.8 |
| Outdoor** | 195 | 32.2 |
| **Monthly income (ETB)** |  |  |
| ≤ 1000 | 172 | 28.4 |
| 1001-2000 | 134 | 22.1 |
| 2001-3635 | 148 | 24.5 |
| ≥ 3636 | 151 | 25.0 |

Where \* (Teacher, office workers, shop keepers, bank accountant)
\*\* (Farmer, construction workers, daily laborers)

Study participants who exposed to dust and wind were 337 (55.7%) and 350 (57.9%) respectively. Participants who exposed to sunlight for more than 5hrs per day were 280 (46.3%) and half of the respondents 302 (49.9%) spend most of their time on outdoor activities. <u>Table 2</u>. From the total study participants only 171 (28.3%) study participants used sunglass or hat. Most of the study participants never smoked cigarettes, no history of dry eye and hypertension with 518 (85.6%), 503 (83.1%) and 558 (92.2%) respectively. <u>Table 3</u>.

**Table 2:** Environmental factors associated with pterygium among adults living in Kolla Diba town, Northwest Ethiopia, June 2019 (n=605)

| Variables | Frequency | Percent (%) |
| --- | --- | --- |
| **Dust exposure** |  |  |
| Yes | 337 | 55.7 |
| No | 268 | 44.3 |
| **Sun exposure** |  |  |
| Non exposed (<5hr) | 325 | 53.7 |
| Exposed (≥5hr) | 280 | 46.3 |
| **Wind exposure** |  |  |
| Yes | 350 | 57.9 |
| No | 255 | 42.1 |
| **Time spent outdoor** |  |  |
| <5hr. | 303 | 50.1 |
| ≥5hr. | 302 | 49.9 |

**Table 3:** Behavioral/ lifestyle factors associated with pterygium among adults living in Kolla Diba town, Northwest Ethiopia, June 2019 (n=605)

| **Variables** | **Frequency** | **Percent (%)** |
| --- | --- | --- |
| **Use of sunglass or hat** |  |  |
| No | 434 | 71.7 |
| Yes | 171 | 28.3 |
| **Drinking status** |  |  |
| Never | 255 | 42.2 |
| Past | 86 | 14.2 |
| Present | 264 | 43.6 |
| **Smoking status** |  |  |
| Never | 518 | 85.6 |
| Past | 64 | 10.6 |
| Present | 23 | 3.8 |
| **Use of traditional eye medication** |  |  |
| No | 514 | 85.0 |
| Yes | 91 | 15.0 |
| **Severe blepharitis** |  |  |
| No | 312 | 51.6 |
| Yes | 293 | 48.4 |
| **History of dry eye** |  |  |
| No | 503 | 83.1 |
| Yes | 102 | 16.9 |
| **History of hypertension** |  |  |
| No | 558 | 92.2 |
| Yes | 47 | 7.8 |
| **Family history of pterygium** |  |  |
| No | 498 | 82.3 |
| Yes | 107 | 17.7 |

The overall prevalence of pterygium among adults living in Kolla Diba town was 112 (18.5% (95% CI (15.6-21.7)). Among those participants, 65 (58%) had unilateral pterygium. Majority of the participants 87 (77.7%) had pterygium on the nasal side and the rest 12 (10.7%) had temporal and 13 (11.6%) had double pterygium. Most of the study participants had grade 1 pterygium, 63 (56.3%) followed by grade 2 pterygium, 38 (33.9%) and the rest grade 3 and 4 accounts less than 8% in total. <u>Table 4</u>.

**Table 4:**
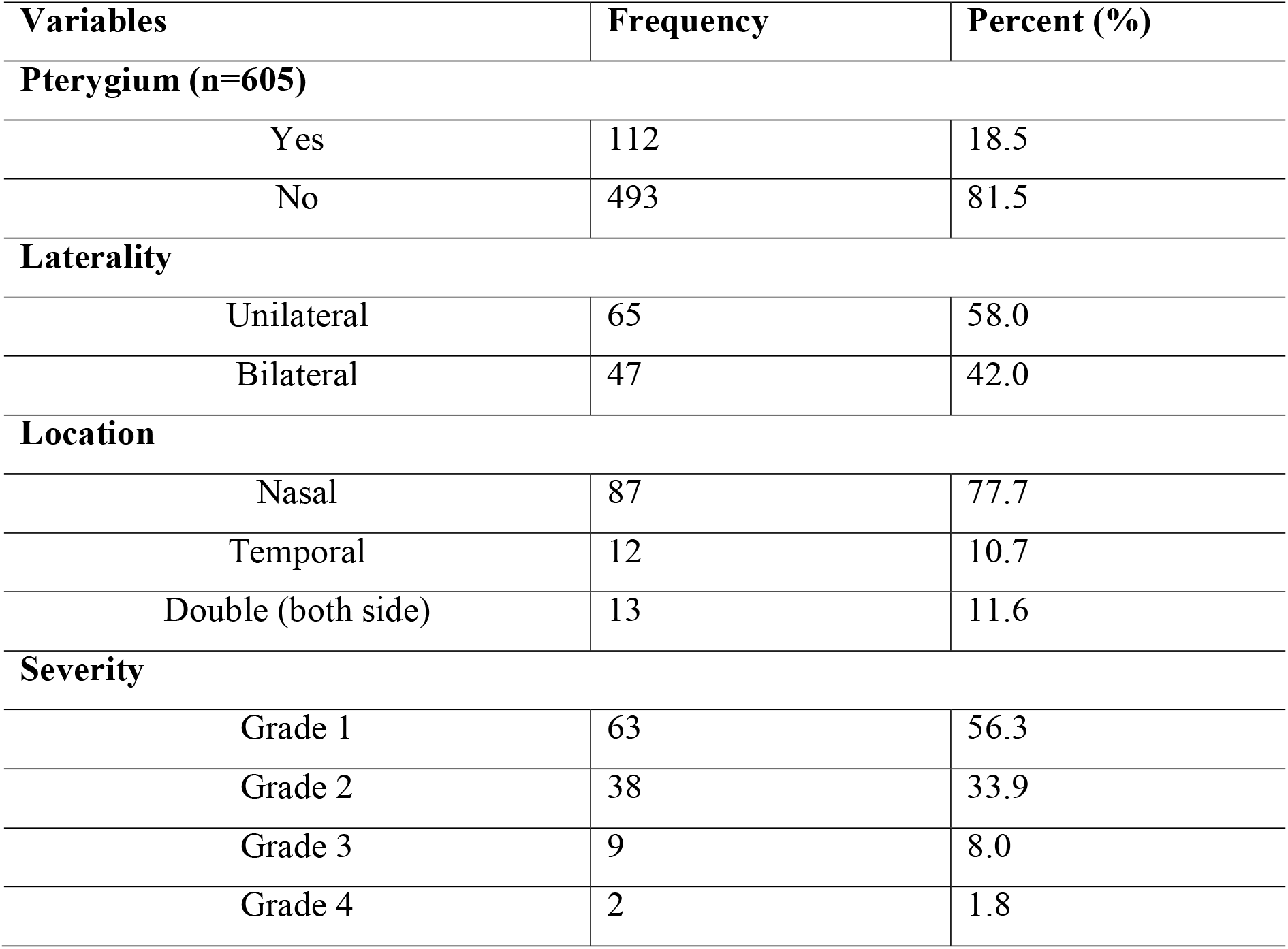
Prevalence, laterality, location and severity of pterygium among adults living in Kolla Diba town, Northwest Ethiopia, 2019

| **Variables** | **Frequency** | **Percent (%)** |
| --- | --- | --- |
| **Pterygium (n=605)** |  |  |
| Yes | 112 | 18.5 |
| No | 493 | 81.5 |
| **Laterality** |  |  |
| Unilateral | 65 | 58.0 |
| Bilateral | 47 | 42.0 |
| **Location** |  |  |
| Nasal | 87 | 77.7 |
| Temporal | 12 | 10.7 |
| Double (both side) | 13 | 11.6 |
| **Severity** |  |  |
| Grade 1 | 63 | 56.3 |
| Grade 2 | 38 | 33.9 |
| Grade 3 | 9 | 8.0 |
| Grade 4 | 2 | 1.8 |

From the Bi-variable logistic regression analysis; factors like age, gender, marital status, educational status, occupation, dust exposure, sun exposure, wind exposure, duration on outdoor activities, use of sunglass or hat, drinking status, smoking status, use of traditional eye medication, severe blepharitis, history of dry eye and history of hypertension were independently associated with pterygium with the significant level of p < 0.20.

In multivariable logistic regression analysis factors such as marital status, occupation, sun exposure, wind exposure, drinking status, and severe blepharitis had statistically significant association with pterygium. This study found that there is a positive association between marital status (being widowed) and the development of pterygium. The odds of developing pterygium among widowed participants were 7 times more likely as compared to single participants (AOR = 7.32 (95%CI: 2.88, 18.57)). Outdoor occupation workers such as farmers, daily laborers, taxi drivers, vendors, construction workers, etc. were 2.50 times smore likely to develop pterygium as compared to indoor workers such as office workers, teachers and so on (AOR = 2.50 (95%CI: 1.46, 4.29)).

Participants who had exposed to wind were 2 times more likely to develop pterygium than non-exposed (AOR = 1.97 (95%CI: 1.04, 3.72)). Alcohol drinking had a positive association with pterygium and those participants who had currently drink alcohol were 2.62 times more likely to develop pterygium as compared to non-drinkers (AOR = 2.26 (95%CI: 1.48, 4.63)). The odds of developing pterygium on study participants who exposed to sun were 2.38 times more likely than non-exposed (AOR = 2.38 (95%CI: 1.28, 4.43)). Those participants who had severe blepharitis were 2.45 times higher risk of developing pterygium (AOR = 2.45 (95%CI: 1.48, 4.05)). <u>Table 5</u>.

**Table 5:** Binary logistic regression of factors associated with pterygium among adults living in Kolla Diba town, Northwest Ethiopia, June 2019(n=605)

| Variables | Pterygium |  | COR(95% CI) | AOR(95% CI ) |
| --- | --- | --- | --- | --- |
|  | Yes | No |  |  |
| Age |  |  |  |  |
| 18-26 | 23 | 146 | 1.00 | 1.00 |
| 27-35 | 22 | 139 | 1.00 (0.54, 1.88) | 0.60 (0.28, 1.30) |
| 36-48 | 34 | 99 | 2.18 (1.21, 3.92) | 0.87 (0.38, 1.96) |
| ≥49 | 33 | 109 | 1.92 (1.07, 3.46) | 0.91 (0.38, 2.15) |
| Gender |  |  |  |  |
| Female | 39 | 249 | 1.00 | 1.00 |
| Male | 73 | 244 | 1.91 (1.25, 2.93) | 1.03 (0.58, 1.83) |
| Marital status |  |  |  |  |
| Single | 20 | 159 | 1.00 | 1.00 |
| Married | 59 | 244 | 1.92 (1.12, 3.31) | 1.44 (0.78, 2.65) |
| Divorced | 19 | 61 | 2.48 (1.24, 4.96) | 1.63 (0.75, 3.57) |
| Widowed | 14 | 29 | 3.84 (1.74, 8.45) | 7.32 (2.88,18.57)*** |
| Educational status |  |  |  |  |
| Unable to read & write | 22 | 77 | 2.53 (1.22, 5.24) | 1.01 (0.38, 2.70) |
| Read & write only | 18 | 62 | 2.57 (1.20, 5.51) | 1.18 (0.47, 3.00) |
| Primary | 22 | 78 | 2.50 (1.20, 5.17) | 1.52 (0.64, 3.61) |
| Secondary | 36 | 152 | 2.10 (1.10, 4.10) | 1.44 (0.67, 3.10) |
| College & above | 14 | 124 | 1.00 | 1.00 |
| Occupation |  |  |  |  |
| Indoor | 41 | 369 | 1.00 | 1.00 |
| Outdoor | 71 | 124 | 5.15 (3.34, 7.96) | 2.50 (1.46, 4.29)*** |
| Dust exposure |  |  |  |  |
| Non exposed | 25 | 243 | 1.00 | 1.00 |
| Exposed | 87 | 250 | 3.38 (2.10, 5.46) | 0.88 (0.42, 1.86) |
| Sun exposure |  |  |  |  |
| Non exposed | 26 | 299 | 1.00 | 1.00 |
| Exposed | 86 | 194 | 5.12 (3.19, 8.24) | 2.38 (1.28, 4.43)** |
| Wind exposure |  |  |  |  |
| No | 19 | 236 | 1.00 | 1.00 |
| Yes | 93 | 257 | 4.50 (2.66, 7.60) | 1.97 (1.04, 3.72)* |
| Duration/ Time spent outdoor |  |  |  |  |
| <5hr | 29 | 274 | 1.00 | 1.00 |
| ≥5hr | 83 | 219 | 3.58 (2.26, 5.66) | 0.75 (0.33, 1.68) |

| Variables | Pterygium |  | COR(95% CI) | AOR(95% CI) |
| --- | --- | --- | --- | --- |
|  | Yes | No |  |  |
| **Use of sunglass or hat** |  |  |  |  |
| Yes | 23 | 148 | 1.00 | 1.00 |
| No | 89 | 345 | 1.66 (1.01, 2.73) | 1.43 (0.82, 2.50) |
| **Drinking status** |  |  |  |  |
| Never | 24 | 231 | 1.00 |  |
| Past | 16 | 70 | 2.20 (1.11, 4.40) | 1.73 (0.81, 3.69) |
| Present | 72 | 192 | 3.61 (2.20, 5.95) | 2.62 (1.48, 4.63)** |
| **Smoking status** |  |  |  |  |
| Never | 84 | 434 | 1.00 | 1.00 |
| Past | 23 | 41 | 2.90 (1.65, 5.10) | 1.26 (0.63, 2.48) |
| Present | 5 | 18 | 1.44 (0.52, 3.97) | 0.70 (0.21, 2.25) |
| **Use of traditional eye medication** |  |  |  |  |
| No | 84 | 430 | 1.00 | 1.00 |
| Yes | 28 | 63 | 2.28 (1.38, 3.76) | 1.12 (0.60, 2.10) |
| **Severe blepharitis** |  |  |  |  |
| No | 30 | 282 | 1.00 | 1.00 |
| Yes | 82 | 211 | 3.65 (2.32, 5.76) | 2.45 (1.48, 4.05)*** |
| **History of dry eye** |  |  |  |  |
| No | 78 | 425 | 1.00 | 1.00 |
| Yes | 34 | 68 | 2.72 (1.69, 4.39) | 0.88 (0.46, 1.67) |
| **History of HTN** |  |  |  |  |
| No | 97 | 461 | 1.00 | 1.00 |
| Yes | 15 | 32 | 2.23 (1.16, 4.27) | 1.47 (0.66, 3.30) |

Where, \* $p \leq 0.05$ ; \*\* $p \leq 0.01$ ; \*\*\* $p \leq 0.001$ ; 1.00 = reference; CI= Confidence Interval

## Discussion

The prevalence of pterygium from various studies in the world shows considerable variation, ranging from 0.07% to 53% depending on the difference in various geographical and environmental factors [2,3,38,39].

In this community-based cross-sectional study the prevalence of pterygium among adults living in Kolla Diba town was 112 (18.5% (95% CI; 15.6–21.7)). This finding is lower as compared to the study conducted, in Gondar city (Ethiopia) 38.7% [2]. This discrepancy might be due to the difference in socioeconomic status and reference age group. In the study conducted in Gondar city 71% of the study participants had low monthly income (< 1000 ETB). But here in this study only 28.4% of the study participants had low monthly income. As previous studies described as low economic status had positively associated with the prevalence of pterygium [11,26,28,37,48]. Another reason might be age cut off point as the present study involved all available inhabitants aged ≥18 years as compared to the study conducted in Gondar city (age >20 years). In contrast to few other studies, this study also shows lower prevalence than other studies conducted in Ghana 31.0% [18], 30.8% in Kumejima (Southwest Japan) [36], 37.46% in Duomen County (Republic of China) [39] and 36.6% in Amazon Basin (Brazil) [31]. This variation might be due to the difference in the target population, ethnicity and altitudinal variation.

This finding is higher when compared it with the study conducted in Meskan district, Southern Ethiopia which was 8.80% [16]. This variation might be due to the difference in target population and ethnicity (the prevalence of pterygium varied among different ethnic groups) [5,9,24,49]. The present study shows higher prevalence of pterygium in comparison to other studies conducted in Saudi Arabia (Alkhobar) 0.07% [38], 13.0% in Rural agrarian central India [22], 10.53% in China (Shandong province) [20], 4.40% in Japan [1], 8.80% in South Korea [26], 2.3% in Russia, 8.12% in Brazil (Sao Paulo (Botucato city)) [32], 11.24% in Hawaii [24], 5.90% in Spain [3], and 1.09% in Portugal [33]. This discrepancy might be due to the difference in geographical location, climatic change, altitudinal variation, life style and other environmental factors. Most of the above studies also done on older adults (40 years and above).

The present study finding is laying in between the highest and the lowest prevalence among different epidemiological studies in the world. This finding is in line with the study conducted in Southeastern Nigeria, Central Myanmar and in Riau Archipelago (tropical islands of Indonesia) which was 19.3%, 19.6% and 17.0% respectively [23,35,50]. This is because Ethiopia, Nigeria, Central Myanmar and Riau Archipelago (Indonesia) located in similar geographical region (pterygium belt tropical regions) in the tropics near the equator. These studies also had similar age group reference and study design.

In many works of literatures marital status had no direct association with pterygium [2,13,28]. But here in this study marital status (being widowed) had a positive association with pterygium. The present study shows widowed participants who had developed pterygium were older in age. The reason may be due to aging since in this study widowed participants were older in age, despite still in debate many authors revealed that pterygium is a degenerative (aging) disorder of the conjunctival tissue that leads to vascular proliferation [2,11,30,49,51]. However; Age had no association with pterygium in this study which is one part of the debate “It is not just a degenerative disease but may be a proliferative disorder of the ocular surface”. Therefore it is due to multifactorial which include genetics (which plays an important role in the pathogenesis) and other environmental factors such as wind, dry atmosphere, dust, chemicals and air pollution may contribute for the development of pterygium [2,7,15,40,52].

Outdoor occupation was significantly associated with the increased prevalence of pterygium. This finding was supported by plenty of previous literature as an outdoor occupation is usually associated with many environmental factors like excessive UV radiation and dust exposure that predisposed to the development of pterygium [15,37,44]. For instance, Nepalese people are equally involved in outdoor activities like farming, animal rearing, and manual labor in order to balance their needs thus increased exposure to potential risk factors like ultraviolet radiation in outdoor jobs [51]. Another study showed that people who spend more than three-quarters of their day outside were more likely to develop pterygium, thus outdoor activity is frequently associated with the degree of UVAF and can damage limbal stem cells by activation of matrix metalloproteinase and leads to pterygium [15,40,45,52].

This study found that there is a positive association between sun exposure and the development of pterygium. This might be due to the fact that prolonged ultraviolet exposure causes biological change in Bowman’s membrane and that altered proteins so formed could then act as an angiogenic or ‘pterygiogenic’ factor [24,28,40]. Many other previous studies also found a strong positive association between sun exposure and pterygium as excessive sun exposure cause damage to limbal stem cells by activation of matrix metalloproteinase which leads to pterygium [24–26,37,40,42].

There was also a positive association between wind exposure and the prevalence of pterygium. This finding is supported by the study conducted in Hawaii, showed that increased risk of exposure to wind while surfing and doing outdoor activities may contribute to pterygium development due to the entrance of airborne debris in to the eye [24].

According to this study, alcohol drinking had a positive association with pterygium. This finding is also supported by previous studies and described as alcohol consumption was another lifestyle-related factor related to low socioeconomic status that leads to the development of pterygium [15,37]. But contrary to this, a study conducted in Xinjiang, China revealed that alcohol consumption had no influence on the development of pterygium [9].

A significant association between severe blepharitis and pterygium were found and different authors reported similar finding. For instance, A study in Israel and others showed that severe blepharitis was significantly associated with pterygium development [13,21,28]. In severe blepharitis, the inflammation and oxidative stress may play a putative role in pterygium pathogenesis by increasing the damage caused by UV radiation on the limbal basal stem cells and accelerating the multistep mutation pathway that leads to pterygium formation. Again in severe blepharitis, there is tear film abnormality which may cause a predisposition to the proliferation of fibro-vascular tissue [28].

Some of the data were subject to recall bias from study participants. This study didn’t conduct laboratory investigations in order to rule out other findings related to pterygium such as OSSN (ocular surface squamous neoplasia] and this may compromise the result.

Being a cross-sectional study was also another limitation. It’s difficult to determine causality between the risk factors and pterygium in this study.

## Conclusion

The prevalence of pterygium in Kolla diba town is about 18.5%, which is relatively higher. Marital status (being widowed], outdoor occupation, alcohol drinking, severe blepharitis, excessive sun exposure and wind exposure had statistical significant associated factors for the development of pterygium.

## Data Availability

All data are fully available without restriction

http://www.PONE-D-19-33031

## Acknowledgement

I would like to acknowledge University of Gondar for all the necessary services like diagnostic materials support to conduct this study.

My appreciation also goes to supervisors, data collectors and study participants for their cooperation in taking part in this study.

